# Knowledge, Attitude, Practices, and Determinants of Them towards Tuberculosis in Bangladesh: A Cross-sectional Study

**DOI:** 10.1101/2022.01.04.21268590

**Authors:** Sultan Mahmud, Md Mohsin, Saddam Hossain Irfan, Abdul Muyeed, Ariful Islam

**Author notes:** Corresponding Author: Md Mohsin.

## Abstract

**Introduction:** Tuberculosis (TB) is an infectious disease that causes thousands of deaths in Bangladesh. Bangladesh is one of the high-risk countries among 30 high TB burden countries. In this study, we aimed to assess the knowledge, practices, attitudes towards TB, and the factors associated with them in the general population of Bangladesh.

**Method:** A web-based anonymous cross-sectional survey was conducted among the general population in Bangladesh. A comprehensive consent statement was included at the beginning of the survey by explaining the study’s intent, types of questions, anonymous and voluntary nature. Analysis was carried out using the chi-square test and univariate and multivariate logistic regression.

**Results:** Among 1,180 eligible respondents, 58.64% were males, and 62.37% were married. The majority of the participants (78.28%) were aged between 15 to 44 years. Overall adequate knowledge, favorable attitudes, and good practices about TB were found respectively in 47.8%, 44.75%, and 31.19% of the general population of Bangladesh. Almost the same sets of associated factors were found to influence adequate knowledge, favorable attitudes, and good practices towards TB among general people. Males, young, unmarried, respondents with higher education, and urban respondents were more likely to have adequate knowledge, favorable attitudes, and good practices towards TB.

**Conclusion:** Policymakers need to design programs and interventions to improve knowledge, attitudes, and good practices towards TB among the general people by focusing on vulnerable groups such as females, young and older people, people who live in the rural areas, and illiterate/less educated people.

**Key Questions:** *What is already known?:*

- TB is highly prevalent in Bangladesh. It is among the 30 most-affected countries globally by TB and carries almost 4% of the global TB burden.
- One of the significant probable obstacles to preventing, controlling, and eliminating tuberculosis is a lack of knowledge and understanding about the disease and a negative attitude toward it. Additionally, there are numerous misconceptions about the etiology and route of transmission of the disease in Bangladesh.

*What are the new findings?:*

- A low level of appropriate knowledge, positive attitudes, and good practices were found in the study.
- Overall, adequate knowledge, favorable attitudes, and good practices about tuberculosis were discovered only in 47.8%, 44.75%, and 31.19%, respectively, of Bangladesh’s general population. Males, those who are young and unmarried, those with a higher level of education, and those who live in urban areas were more likely to have enough knowledge, favorable attitudes, and appropriate practices about tuberculosis.

*What do the new findings imply?:*

- Bangladesh showed little progress in fighting against TB in the recent past. Therefore, government and policymakers should renew their efforts to improve knowledge, attitudes, and practices as an essential step to achieve stipulated TB goals.
- Further studies are required that also include TB patients, healthcare workers, and community influencers.

## 1 Introduction

Tuberculosis (TB) is an infectious bacterial disease caused by Mycobacterium tuberculosis (MTB) [1]. TB remains a severe health problem worldwide, despite a tremendous performance in controlling the disease, with an estimated 37 million lives saved by improved diagnosis and treatment since 2000 [2]. Per a 2015 global health report, tuberculosis (TB) is the leading cause of morbidity and mortality globally, ranking alongside the human immunodeficiency virus (HIV) [3]. Globally, 10.4 million people were reported to have contracted tuberculosis in 2015, with 1.8 million people dying from the disease [4]. However, developing countries bear the brunt of tuberculosis’s impact. In 2015, almost 95% of the estimated 1.8 million TB deaths occurred in low- and middle-income countries [3].

Globally, 10 million individuals were infected with tuberculosis in 2019, with 79% of those infected living in the 30 high-burden countries and 1.2 million people dying from the disease [5]. Bangladesh is one of the 30 countries with the highest TB burden, accounting for 3.6 percent of the global total [6]. In Bangladesh, the estimated incidence of tuberculosis per 100,000 is 221, with a death rate of 24 per 100,000 [5]. According to the Global TB Report 2020, 0.7% of new cases and 11% of previously treated patients in Bangladesh are positive for multidrug-resistant tuberculosis (MDR-TB), which has an incidence rate of 2.0 per 100,000 people [5].

Even though tuberculosis is a preventable and treatable disease, the situation in Bangladesh has remained essentially constant over the years, with moderate progress and no signs of a breakthrough in the near future [6]. Bangladesh established End TB goals, including a 95% reduction in TB mortality and a 90% reduction in TB incidence by 2035 compared to 2015 levels, with intermediary goals set for 2020, 2025, and 2030 [7]. The Stop TB Partnership has issued TB diagnosis and treatment targets for Bangladesh for 2018–2022 as a result of the United Nations High-Level TB Meeting (UNHLM). To meet the UNHLM’s cumulative five-year TB targets, Bangladesh must raise diagnosis and treatment by 45 percent above the total notifications reported during the pre-UNHLM five-year period (2014–2017) [8].

The National Tuberculosis Control Program (NTP) has chosen the Directly Observed Treatment, Short-course (DOTS) technique to lessen this burden, which is predominantly given through government-run health institutions [9]. However, considerable impediments to implementation exist, mainly due to insufficient infrastructure and suitable health workers [10]. Therefore, the World Health Organization recommends that national TB Control Programs use an Advocacy, Communication, and Social Mobilization (ACSM) framework to address these issues. This strategy framework targets four significant issues: enhancing case detection and treatment adherence, eliminating stigma and prejudice, empowering tuberculosis patients, and mobilizing the resources and political commitment needed to combat the disease [11].

Despite the efforts, the expected degree of improvements in controlling the TB crisis has not been made yet in Bangladesh [12]. One of the primary challenges in preventing, controlling, and eliminating tuberculosis is a lack of awareness and knowledge and a negative attitude about the disease [1]. There are also a lot of misconceptions concerning the etiology and mode of transmission in Bangladesh [13]. TB is thought to be inherited in some locations [14–16]. A lack of understanding about tuberculosis and old misconceptions are linked to delays in case detection and treatment for TB [15,17]. The widespread prejudice towards TB/HIV patients, misapprehension of transmission of TB and other infectious diseases, and poor knowledge about treatment of infectious diseases are the serious restrictions in achieving millennium development goals related to TB and other infectious diseases [18–20].

The authors of this study understand that the lack of knowledge, misconceptions and bad practices among general people could be the reasons for slow and unsatisfactory progress in the fight against TB in Bangladesh. Unfortunately, there is a lack of large-scale and quality studies in Bangladesh that explore the knowledge and attitude about TB and practices to prevent it. Therefore, this study aimed to investigate general people’s knowledge, attitude, and practices towards TB. Also, this study explores the risk factors associated with poor knowledge, attitude, and practices towards TB among general people. The government should renew their commitment to national tuberculosis control activities based on data-driven, effective methods to meet the stipulated goals. The findings of this study would be a significant help for the government and policymakers in this regard.

## 2 Methods

### 2.1 Study Design and study participants

This study was a cross-sectional approach to collect data regarding knowledge, attitude, and practices about tuberculosis in Bangladesh. It was an online anonymous, self-interviewed survey conducted from May 20 to August 10, 2021. People aged 15 and over and living in Bangladesh were eligible to participate in this survey. In the beginning, there was a section describing the study’s objective, the idea about the questionnaire, assurance about the respondents’ confidentiality, and the study’s voluntary nature. It was also mentioned that participants could skip a question if it seems sensitive. The online survey started with the respondents’ agreement and the eligibility check. The voluntary participants were also requested to share the survey link with their connections after completion. The study questionnaire was developed in English (S1 Questionnaire), further translated in Bangla. The questionnaire was validated by several experts and pilot surveys. An online survey link (KoBoToolbox) was shared with almost 4000 people of Bangladesh through social media (FB, Messenger, WhatsApp, and Electronic Email). A total of 1,205 (response rate was 30%) people filled up and submitted their responses; among them 25 of the respondents were not eligible (either aged less than 15 or living outside of Bangladesh) for this study.

### 2.2 Sample size

In this study, we aimed to examine the knowledge, practices and attitude towards TB and their associated factors among the general population in Bangladesh. We did not find previous literature from Bangladesh that examined the knowledge, practices, and attitude towards TB and their associated factors among the general population. For calculating desirable sample size, we assume that 50% of the general population have adequate knowledge about TB, good practices and a favorable attitude towards TB. Using an online sample size calculator [21], we found that this study requires a sample size of 1,163 to represent a population size of 164,689,383 [22] with 5% absolute precision, 95% confidence, and an expected response rate of 33%.

### 2.3 Instruments

Respondents were reached through large and popular Facebook groups, Messenger, WhatsApp, and electronic mail of the different groups of respondents by circulating KoBoToolbox online survey link. The structured questionnaire was made of 3 main sections: (i) Background characteristics of the respondents; (ii) Risk behaviors related to tuberculosis; (iii) Knowledge of tuberculosis; (iv) Attitudes towards tuberculosis; and (v) Practice of tuberculosis.

#### 2.3.1 Background Characteristics

At the outset of the survey, we checked the aptness of the participants by asking two questions, “How old are you (in years)?” and “Do you currently live in Bangladesh?” We also added respondents’ socio-demographic and some personal details in this section. The socio-demographic details were gender, current marital status, religion, educational qualification, and socioeconomic details were monthly household income level, occupational status, residence, etc. The second part of the survey questionnaire contained questions linked to risk behaviors related to tuberculosis, including diabetes status, smoking status, the status of drinking alcohol in the last three months, the status of exposure to indoor cooking smoke etc. Demographic covariates of this study were categorized in the following way: Residence: Rural, Urban; and Religion: Muslims, Hindu, Buddhists/Cristian; Sex: Female, Male; Age (year): 15-29, 30-44, 45-59, 60-74, 75+; Marital Status: Married, Unmarried, Others (Divorced, Widowed, Separated); Education: Less or equal SSC (10th grade), HSC (12th grade), undergraduate, Master’s or higher, Never been to school; Last month income (Taka): Less than 10 thousand, 11-20 thousand, 21-30 thousand, 31-40 thousand, greater than 40 thousand, No income; Occupation: Business, Housewife, Govt. employee, Non-govt. employee, Unemployed, Self-employed, Student.

#### 2.3.2 Knowledge about Tuberculosis

In this segment, to measure the level of knowledge concerning tuberculosis, respondents were asked a series of questions under a few sub-segments, “Source of knowledge on TB”, “Knowledge about TB causes”, “Knowledge about the transmission of TB”, “Knowledge about symptoms of TB”, and “Knowledge about availability of TB treatment”. The source of knowledge on TB was assessed by asking two questions, “Did you hear anything about TB?” and “From where and whom did you learn about TB?”. Knowledge about TB causes was evaluated by asking, “What is the major cause of TB?”. Knowledge about the transmission of TB was measured asking the questions, “Is TB spread from person to person through the air when coughing or sneezing?”, “Can TB be transmitted by sharing utensils?”, “Can TB be transmitted through food?”, “Can TB be transmitted through sexual contact?”, and “What is the most common site for TB infection in the body?”. Knowledge about symptoms of TB was assessed asking, “A person who is infected with TB coughs for several (more than 3) weeks?”, “A person who is infected with TB has a persistent fever?”, “A person who is infected with TB sweats during the night?”, “A person who is infected with TB has pain in the chest or back?”, and “Weight loss is one of the symptoms of TB?”. Knowledge about TB treatment availability was assessed by asking, “Is TB management available free of cost in Bangladesh?” and “Is TB curable?”

#### 2.3.3 Attitudes and Practices about Tuberculosis

In this section, they were asked a series of questions to assess the level of attitude and practices regarding tuberculosis. Attitudes regarding TB were assessed asking the questions “In your opinion, how serious a disease is TB?”, “Are you afraid of getting infected with TB?”, “Will you keep secret when any family member gets TB?”, “Would you be willing to work with someone previously treated for TB?” and “What would be your reaction if you were found out that you have TB?”. Practices regarding TB among respondents were measured by asking two questions, “If you had symptoms of TB, at what point would you go to the health facility?” and “If you had symptoms of TB, where will you go for TB treatment?

### 2.4 Consent and Ethical Considerations

At the outset of the survey, a section described the study’s eligibilty, aims, the questionnaire’s concept, assurances regarding respondents’ confidentiality, and the study’s voluntary nature. Additionally, it was indicated that participants could omit a question if it appeared to be sensitive. This study was reviewed and waived the requirement of an IRB approval by the Ethical Review Committee, Faculty of Biological Science and Technology, University of Science and Technology, Jashore, Bangladesh. Because this was an anonymous online survey, it was voluntary, and it did not include any clinical operations.

### 2.5 Statistical analysis

This study’s primary outcome of interest was respondents’ adequate knowledge, good practices, and positive attitudes towards TB. Participants’ knowledge of the cause, mode of transmission, signs and symptoms and treatment availability of TB was coded as “1” and labeled as “adequate knowledge” if the respondent correctly answered ≥ 50% (≥ 7 questions out of the total 14) of questions. Otherwise, participants’ knowledge was coded as “0” and labeled as “poor knowledge”. The overall participents’ attitude towards TB was defined as “Favorable attitude” and coded as “1” if the respondent correctly answered ≥ 3 questions out of the total 5 and otherwise defined “Unfavorable attitude” and coded as “0”. We used two questions to assess respondents’ practice towards TB (Q1: If you had symptoms of TB, at what point would you go to the health facility? and Q2: If you had symptoms of TB, where will you go for TB treatment?). The overall respondents’ practices towards TB were defined as “Good practice” if the respondent correctly answered both questions otherwise define “Bad practice”.

The exploratory analysis (frequencies analysis, means, median, bivariate analysis) was done to check socio-demographic characteristics. The statistical significance of the correlation between socio-demographic factors and knowledge of the respondents and their practice and attitude towards TB was inspected using the Chi-square test. All the significant factors at a 10% level of significance in the Chi-square test were included in the univariate logistic regressions [23]. We did so to recheck the association between socio-demographic factors and knowledge of the respondents and their practice and attitude towards TB. The adjusted odds ratios (AOR) were also calculated using multivariate logistic regressions [24,25] with a 95% confidence interval (CI). All the analyses were done by using the Statistical package STATA version 16.0.

### 2.6 Patient and Public Involvement

This study did not include any patients. It was an online-based, voluntary, and anonymous study that collected data from general people aged 15 years or over and living in Bangladesh. A comprehensive consent statement was included at the beginning of the survey describing the study’s objectives, nature, types of questions to be asked, skipping options, etc. The consent also assured that the data will be used in a combined form only for research purposes.

## 3 Results

### 3.1 Socio-demographic characteristics

More than four thousand people were invited to participate in the survey through online platforms (WhatsApp, Messenger, Email, Linkedin, etc.). A total of 1,180 (30% response rate) people submitted the self-consent completed surveys. The socio-demographic characteristics of the respondents are presented in Table 1. The majority of the respondent tended to be male (58.64%), aged between 15 to 44 years (78.28%), and married (62.37%). Less than one-third of the respondents (34.8%) had a Master’s or higher degree. Most of the respondents were Muslim (87.12%), living in rural areas (56.61%). Almost half of the respondents were students (44.75%).

**Table 1:** Distribution of socio-demographic characteristics of respondents.

| Varibale | Labels | N (%) |
| --- | --- | --- |
| Consent | Yes | 1,180 (100.00) |
| Gender | Male | 692 (58.64) |
|  | Female | 488 (41.36) |
| Age | 15-29 | 404 (34.24) |
|  | 30-44 | 520 (44.07) |
|  | 45-59 | 240 (20.34) |
|  | 75+ | 16 (1.36) |
| Marital status | Married | 736 (62.37) |
|  | Unmarried | 328 (27.80) |
|  | Divorced/Widowed/Separated | 116 (9.83) |
| Education | Less or equal HSC (<=12<sup>th</sup> grade) | 332 (28.14) |
|  | Undergraduate | 408 (34.58) |
|  | Master's or higher (Graduate) | 440 (37.29) |
| Income | Less than 30,000 | 276 (23.39) |
|  | 30,000-45,000 | 176 (14.92) |
|  | 46,000-60,000 | 160 (13.56) |
|  | 61,000-75,000 | 280 (23.73) |
|  | 76,000 and above | 288 (24.41) |
| Occupation | Service holder (govt/private) | 324 (27.46) |
|  | Entrepreneur/business | 172 (14.58) |
|  | Student | 528 (44.75) |
|  | Housewife/Retired/Unemployed/Other | 156 (13.22) |
| Religion | Islam | 1,028 (87.12) |
|  | Hinduism | 134 (11.36) |
|  | Buddhists/Cristian | 18 (1.53) |
| Region | Urban | 512 (43.39) |
|  | Rural | 668 (56.61) |

### 3.2 Knowledge about tuberculosis and associated factors

All the respondents confirmed that they heard about TB. In response to a multiple responses question, we found that the source of information about TB for 85.76% of the respondents was TV/Radio/Newspaper (Figure 1). The second major source was leaflets/Poster/Signboard/ Billboard (66.78%). Nearly half of the respondents received information from health professionals, and 44.4% received it from the internet. Religious leaders/teachers were the sources of information for 43.05% of the respondents. A similar proportion of the participants (41.36%) learned about TB from friends/relatives/family members. Exposure to TB treatment and inmates suffering from TB were the sources of TB for 27.8% and 17.29% of the participants. Table 2 shows that a large proportion (71%) of the participants knew that TB germ or Bacteria is the major cause of TB. Nearly 42% of the respondents knew the correct transmission mode of TB (TB can be spread from person to person through the air when coughing or sneezing).

**Table 2:** Knowledge of respondents on TB cause, transmission, sign & symptom, treatment and attitude and practices towards TB in the general population of Bangladesh.

| Question | Label | N(%) |
| --- | --- | --- |
| **Knowledge about TB causes** |  |  |
| What is the primary cause of TB? | TB germ /Bacteria | 828 (70.41) |
|  | Virus | 188 (15.99) |
|  | Cold wind | 8 (0.68) |
|  | Smoking | 36 (3.06) |
|  | Spoiled soil (soil with a bad odor) | 0 (0) |
|  | Poor hygiene Alcohol | 4 (0.34) |
|  | Inherited | 0 (0) |
|  | Don't know | 112 (9.52) |
| **Knowledge about the transmission of TB** |  |  |
| TB is spread from person to person through the air when coughing or sneezing? | Yes | 488 (41.36) |
|  | No | 672 (56.95) |
|  | Don't know | 20 (1.69) |
| TB can be transmitted by sharing utensils? | Yes | 584 (49.49) |
|  | No | 556 (47.12) |
|  | Don't know | 40 (3.39) |
| TB can be transmitted through food? | Yes | 572 (48.47) |
|  | No | 600 (50.85) |
|  | Don't know | 8 (0.68) |
| TB can be transmitted through sexual contact? | Yes | 624 (52.88) |
|  | No | 532 (45.08) |
|  | Don't know | 24 (2.03) |
| What is the most common site for TB infection in the body?<br>(Only one answer) | Lungs | 952 (80.68) |
|  | Glands | 140 (11.86) |
|  | Brain | 0 (0) |
|  | Bones | 4 (0.34) |
|  | Others (specify) --- | 0 (0) |
|  | Don't know | 84 (7.12) |
| **Knowledge about symptoms of TB** |  |  |
| A person who is infected with TB coughs for several (more than 3) weeks? | Yes | 488 (41.36) |
|  | No | 684 (57.97) |
|  | Don't know | 8 (0.68) |
| A person who is infected with TB has persistent fever? | Yes | 404 (36.33) |
|  | No | 700 (62.95) |
|  | Don't know | 8 (0.72) |
| A person who is infected with TB sweats during the night? | Yes | 512 (43.39) |
|  | No | 652 (55.25) |
|  | Don't know | 16 (1.36) |
| A person who is infected with TB has pain in the chest or back? | Yes | 512 (43.39) |
|  | No | 664 (56.27) |
|  | Don't know | 4 (0.34) |
| Weight loss is one of the symptoms of TB? | Yes | 492 (41.69) |
|  | No | 668 (56.61) |
|  | Don't know | 20 (1.69) |
| **Knowledge about availability of TB treatment** |  |  |
| Is TB management available free of cost in Bangladesh? | Yes | 872 (73.9) |
|  | No | 176 (14.92) |
|  | Don't know | 132 (11.19) |
| Is TB curable? | Yes | 1020 (86.44) |
|  | No | 140 (11.86) |
|  | Don't know | 20 (1.69) |
| **Attitude towards TB** |  |  |
| In your opinion, how serious disease is TB? | Very serious | 936 (79.32) |
|  | Somewhat serious | 112 (9.49) |
|  | Not very serious | 132 (11.19) |
| Do you afraid to get infected with TB? (chose only one) | Yes | 544 (46.1) |
|  | No | 624 (52.88) |
|  | Don't know | 12 (1.02) |
| Will you Keep it secret when any family member gets TB? | Yes | 976 (82.71) |
|  | No | 196 (16.61) |
|  | Don't know | 8 (0.68) |
| Would you be willing to work with someone previously treated for TB? | Yes | 528 (44.75) |
|  | No | 648 (54.92) |
|  | Don't know | 4 (0.34) |
| What would be your reaction if you were found out that you have TB? (chose only one) | Go to pharmacy | 536 (45.42) |
|  | Go to a health facility | 448 (37.97) |
|  | Got to traditional healer | 144 (12.2) |
|  | Pursue other self-treatment options (herbs, etc.) | 44 (3.73) |
|  | Others (specify) | 8 (0.68) |
| **Practice towards TB** |  |  |
| If you had symptoms of TB, at what point would you go to the health facility? (choose only one) | When treatment on my own does not work. | 208 (17.63) |
|  | When symptoms that look like TB signs last for 3–4 weeks. | 336 (28.47) |
|  | As soon as I realize that my symptoms might be related to TB. | 568 (48.14) |
|  | I would not go to the doctor. | 68 (5.76) |
| If you had symptoms of TB, where would you go for TB treatment? (chose only one) | Modern drugs | 832 (70.51) |
|  | Herbal Remedies | 148 (12.54) |
|  | Home Remedies | 124 (10.51) |
|  | Praying /holy water | 44 (3.73) |
|  | Don't Know | 32 (2.71) |

**Figure 1:**
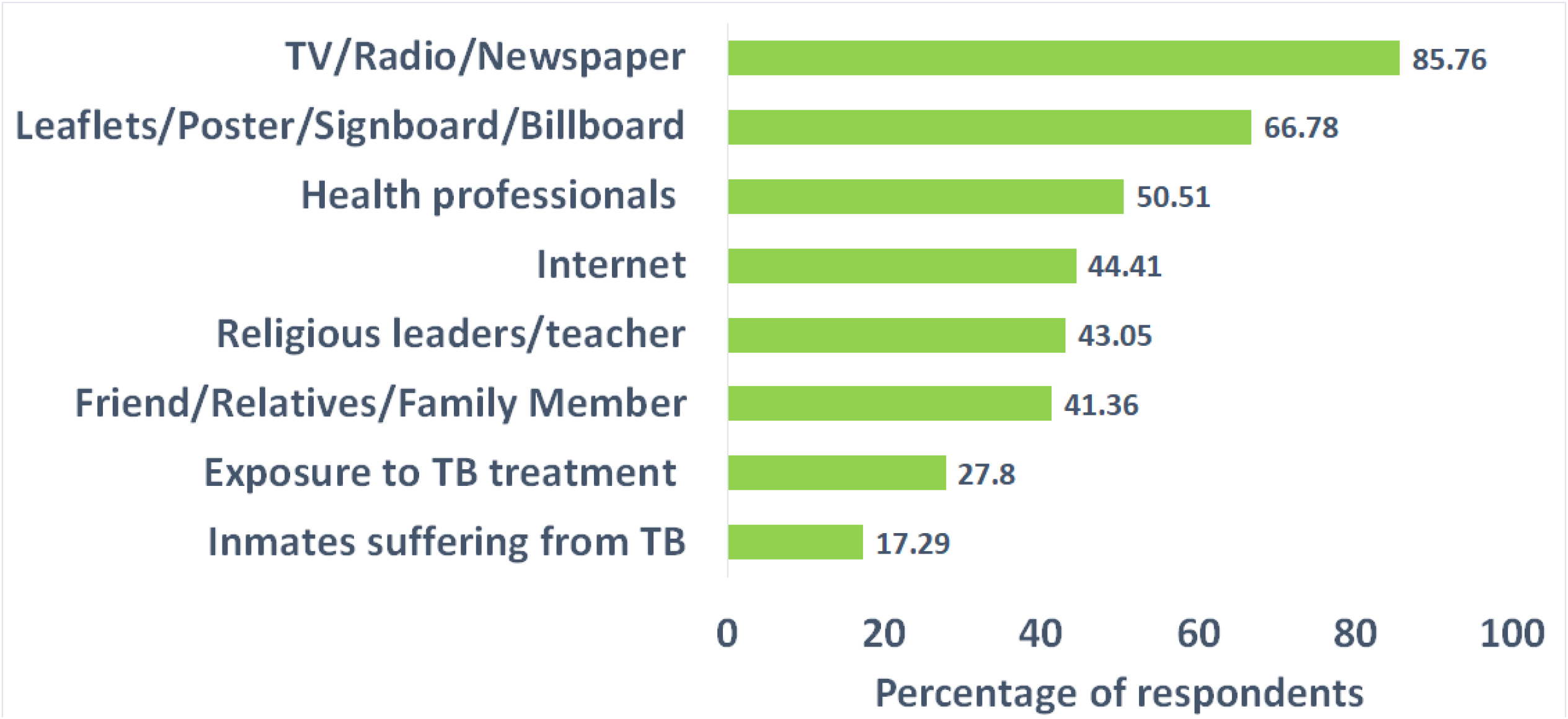
Source of information of the respondents about TB.

Nevertheless, the misconception was observed among a considerable proportion (57%) of the respondents. Almost half of the respondents knew that TB could be transmitted from person to person by sharing utensils and food or by sexual contact. A more significant proportion (81%) of the respondents correctly knew that the lung is the most common site for TB infection in the body. However, the respondent’s knowledge about symptoms of TB was deficient. Out of 1,180 persons, 652 (55.25%) did not know that cough for several (more than 3) weeks is a common symptom of TB infection. Less than 50% (36.33%, 43.39%, and 41.69%, respectively) of the respondents knew that persistent fever, sweats during the night, and weight loss are TB symptoms. Knowledge about the availability of TB treatment was considerably high among the respondents. Almost 85% of the respondents knew that TB is curable, and 74% knew that TB treatment is available and accessible in Bangladesh.

We observed adequate overall knowledge regarding TB in only 47.8% of the respondents (Figure 2). In Appendix A Table 1 shows that 68.21% of males and only 18.85% of females had adequate knowledge about TB. The findings from multivariate regression (presented in Table 3) also show that females had a 90% lower chance of having adequate knowledge about TB than their male counterparts. The age of the respondents was also a significantly associated factor for having adequate knowledge regarding TB. Middle-age people were more likely to have more knowledge about TB. More explicitly, respondents aged 30-40 had a 2.13 times higher likelihood of having adequate knowledge than respondents aged 15-29. However, older respondents (aged 45-59) had around 84% lower chance of having adequate knowledge about TB than the young respondents (aged 15-29). Marital status, education level, and income were strongly correlated with the overall level of knowledge. Around 42% of married and 75.61% of unmarried respondents had adequate knowledge about TB (in Appendix A Table 1). The odds of having adequate knowledge among the unmarried respondents were four times higher than the odds of having adequate knowledge among the married respondents (Table 3). Among respondents who completed undergraduate or running, 62.75% had adequate knowledge, and among respondents with master’s or higher degree, 58.18% had adequate knowledge. However, only 15.66% of the respondents with less or equal HSC degrees (<=12 grade) had adequate knowledge (in Appendix A Table 1). The respondents with education level undergraduate and graduate had respectively 5 and 3 times higher chance of having adequate knowledge than respondents with an education level less or equal HSC (Table 3). Respondents having higher income were more likely to have adequate knowledge about TB. People living in urban areas were more likely to have adequate knowledge. The respondents who live in rural areas had a 69% lower chance of having adequate TB knowledge.

**Table 3:** Associated factors with knowledge of TB among the general population.

| Factors |  | Bivariate analysis |  | Multivariate analysis |  |
| --- | --- | --- | --- | --- | --- |
|  |  | UOR (95% CI) | P-value | AOR (95% CI) | P-value |
| Gender | Male | Ref |  | Ref |  |
|  | Female | 0.11 (0.08-0.14) | <.001 | 0.1 (0.07-0.15) | <.001 |
| Age | 15-29 | Ref |  | Ref |  |
|  | 30-44 | 2.11 (1.62-2.75) | <.001 | 2.13 (1.37-3.32) | <.01 |
|  | 45-59 | 0.27 (0.18-0.39) | <.001 | 0.16 (0.08-0.31) | <.001 |
|  | 75+ | 0.4 (0.13-1.26) | NS | 1.94 (0.19-19.45) | NS |
| Marital Status | Married | Ref |  | Ref |  |
|  | Unmarried | 4.31 (3.22-5.77) | <.001 | 4.01 (2.51-6.42) | <.001 |
|  | Other<sup>f</sup> | 0.1 (0.05-0.21) | <.001 | 0.02 (0-0.06) | <.001 |
| Education | Less or equal HSC | Ref |  | Ref |  |
|  | Graduate | 9.07 (6.34-12.97) | <.001 | 5.06 (2.93-8.74) | <.001 |
|  | Master or higher | 7.49 (5.27-10.65) | <.001 | 3.71 (2.2-6.27) | <.001 |
| Income | Less than 30,000 | Ref |  | Ref |  |
|  | 30,000-45,000 | 0.4 (0.26-0.62) | <.001 | 0.27 (0.14-0.53) | <.001 |
|  | 46,000-60,000 | 0.67 (0.44-1.01) | NS | 0.35 (0.18-0.69) | <.01 |
|  | 61,000-75,000 | 1.39 (0.99-1.94) | NS | 0.94 (0.54-1.65) | NS |
|  | 76,000 and above | 7.78 (5.25-11.52) | <.001 | 9.12 (4.35-19.11) | <.001 |
| Occupation |  |  |  |  |  |
|  | Service holder (govt/private) | Ref |  | Ref |  |
|  | Entrepreneur/business | 0.8 (0.54-1.18) | NS | 0.64 (0.33-1.25) | NS |
|  | Student | 0.22 (0.16-0.3) | <.001 | 0.09 (0.05-0.17) | <.001 |
|  | Other<sup>‡</sup> | 0.37 (0.25-0.54) | <.001 | 0.21 (0.11-0.42) | <.001 |
| Religion | Islam | Ref |  | Ref |  |
|  | Hinduism | 2.14 (1.48-3.12) | <.001 | 2.54 (1.36-4.74) | <.01 |
|  | Buddhists/Cristian | 1.5 (0.59-3.82) | NS | 2.37 (0.4-14.02) | NS |
| Region | Urban | Ref |  | Ref |  |
|  | Rural | 0.58 (0.46-0.73) | <.001 | 0.31 (0.2-0.48) | <.001 |

NS, not significant; Other<sup>‡</sup> includes Housewife, Retired, and Unemployed; Other<sup>£</sup> includes Divorced, Widowed, and Separated

**Figure 2:**
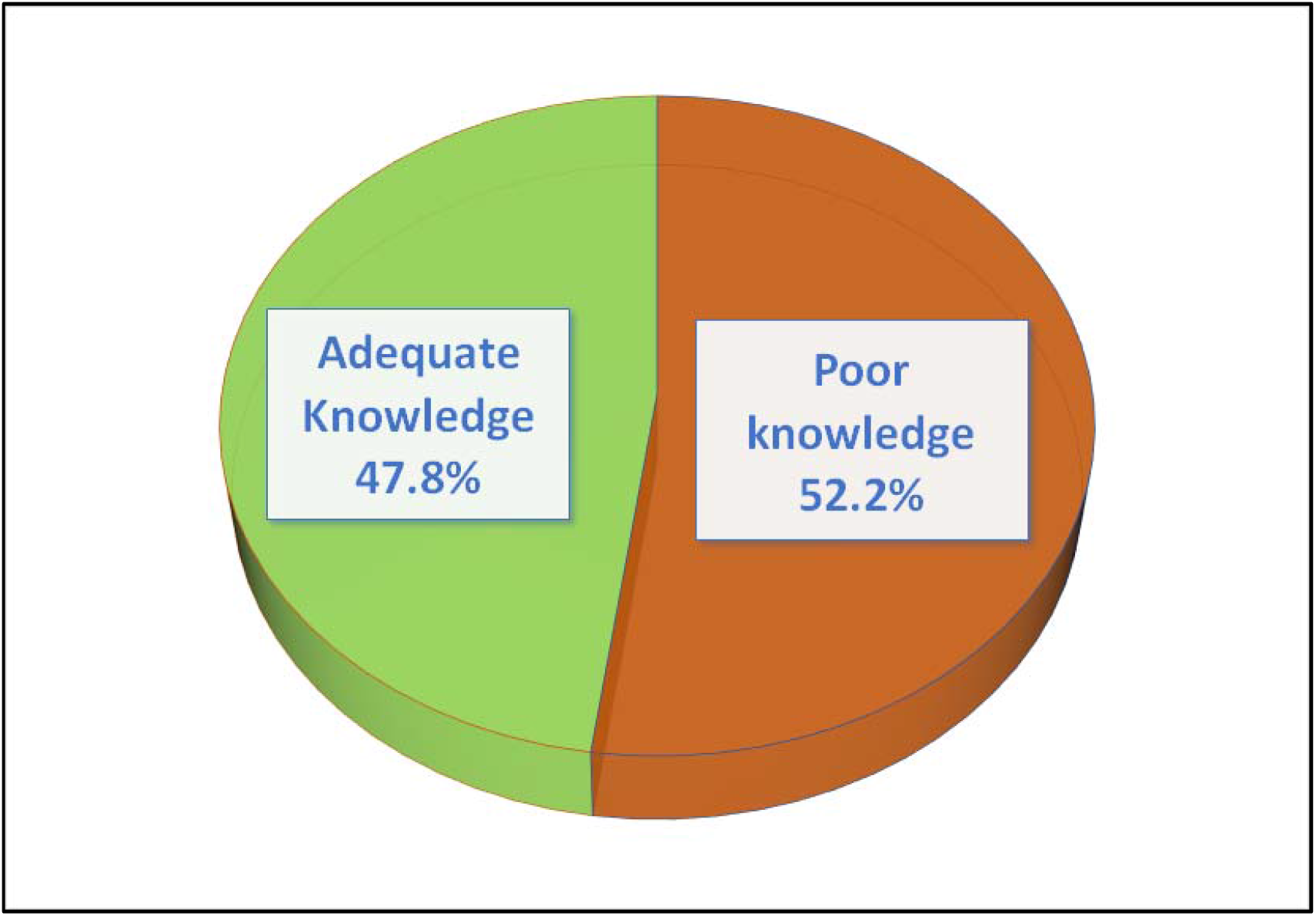
Distribution of knowledge about TB among the general population.

### 3.3 Practices towards tuberculosis and associated factors

Nearly half of the participants preferred to visit health care centers (48.14%) as soon as they realized that their symptoms might be related to TB and wanted to take modern drugs (71%) (Table 2). Less than one-third of 1,180 respondents (31.19%) showed overall good practices (Figure 3). According to bivariate analysis (see Appendix A Table 1), 37.57% of males and 22.13% of females were doing good practices. According to regression findings, female respondents had a 41% lower chance of doing good practices towards TB than males (Table 4). The respondents aged 30-44 had a 1.42 times higher chance of doing good practices than younger respondents (aged 15-29). Good practices were also observed among 28.26% of married and 41.46% of unmarried respondents (Appendix A Table 1). Unmarried respondents had a 1.46 times higher likelihood of having good practices than married respondents (Table 4). Among respondents with education level undergraduate and graduate, 30% had good practices while 21.69% of respondents with less or equal HSC degrees (<=12 grade) had good practices. The odds of having good practices among the respondents with a graduate-level education were two times higher than respondents with less or equal HSC degrees (Table 4). Among the respondents who believe in Hinduism, 55.22% had good practices towards TB and those who believe in Islam, 28.21% had good practices (Appendix A Table 1). The odds of having good practices towards TB among students and entrepreneurs/businesses, respectively, were 1.45 and 1.93 times higher than the odds of having good practices among service holders (govt/private).

**Table 4:**
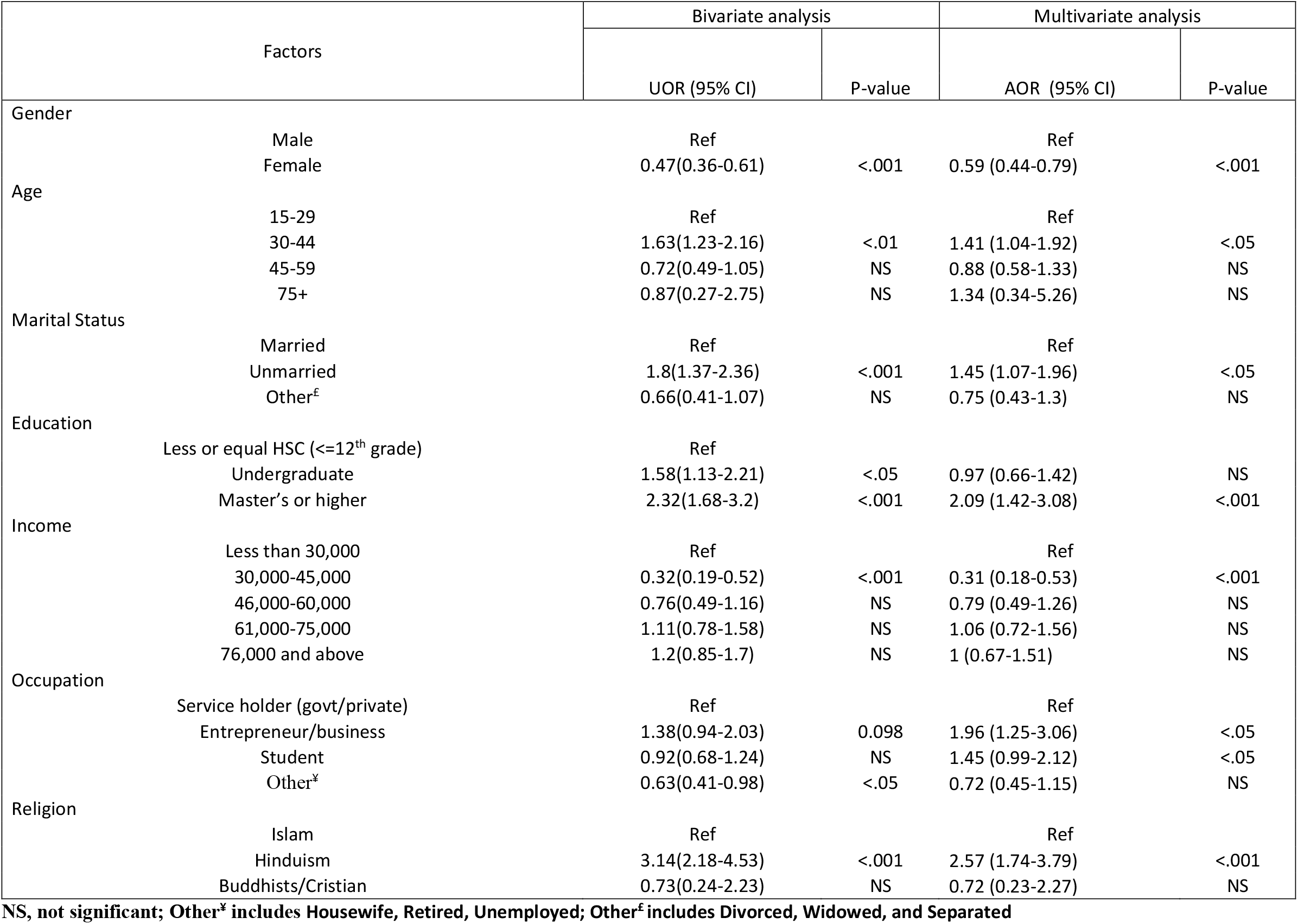
Associated factors with practice towards TB among the general population.

**Figure 3:**
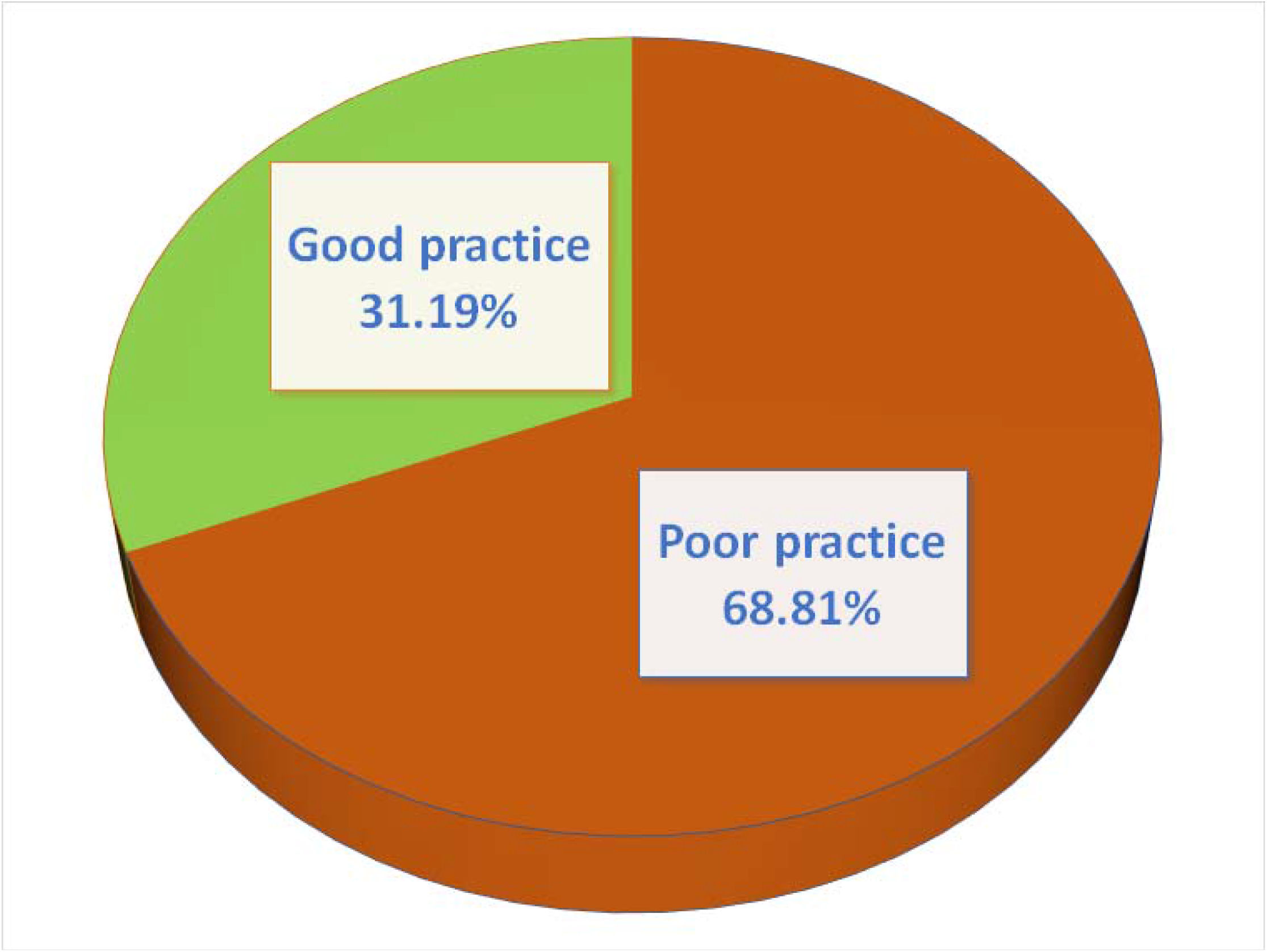
Distribution of practices towards TB among the general population.

### 3.4 Attitude towards tuberculosis and associated factors

According to 79.32% (936) of the respondents, TB is a severe disease (Table 2). Almost half of the participants were afraid to get infected with TB. A large proportion (83%) of the respondents wanted to keep it secret when any family member gets TB. A significant portion of the respondents (54.92%) were unwilling to work with someone previously treated for TB. Also, a considerable proportion (44.75%) of the respondents had stigmatizing thoughts towards TB patients. Figure 4 depicts that almost 45% of the respondents expressed a favorable attitude towards TB. The general population’s attitude in Bangladesh towards TB and associated factors are shown in Table 5. Gender was one of the significant factors of favorable attitudes towards TB. The odds of having a favorable attitude towards TB among females were 95% lower than the odds of having a favorable attitude towards TB among males. The age of the respondents is also a significantly associated factor for having a favorable attitude towards TB. Middle-age people were more likely to have a favorable attitude towards TB. More explicitly, respondents aged 30-40 had a 3.78 times higher likelihood of having a favorable attitude towards TB than respondents aged 15-29. However, older (45-59) respondents had around 69% lower chance of having a favorable attitude towards TB than the young respondents (aged 15-29). Marital status, education level, and income were highly correlated with the overall attitude of respondents towards TB. The odds of having a favorable attitude towards TB among the unmarried respondents were three times higher than the odds of having a favorable attitude towards TB among the married respondents. The respondents with undergraduate and graduate-level education had respectively 6 and 2 times higher chance of having a favorable attitude towards TB than respondents with education level less or equal HSC (<=12^th^ grade). The respondents who live in rural areas had a 54% lower chance of having a favorable attitude towards TB.

**Table 5:** Associated factors with attitude towards TB among the general population.

| Factors |  | Bivariate analysis |  | Multivariate analysis |  |
| --- | --- | --- | --- | --- | --- |
|  |  | UOR (95% CI) | P-value | AOR (95% CI) | P-value |
| Gender | Male | Ref |  | Ref |  |
|  | Female | 0.07 (0.05-0.1) | <.001 | 0.05 (0.03-0.07) | <.001 |
| Age | 15-29 | Ref |  | Ref |  |
|  | 30-44 | 2.93 (2.24-3.83) | <.001 | 3.78 (2.44-5.86) | <.001 |
|  | 45-59 | 0.37 (0.25-0.55) | <.001 | 0.31 (0.16-0.6) | <.001 |
|  | 75+ | 1 (0-0) | NA | 1 (0-0) | NA |
| Marital Status | Married | Ref |  | Ref |  |
|  | Unmarried | 3.94 (2.97-5.21) | <.001 | 3.02 (1.93-4.73) | <.001 |
|  | Other<sup>f</sup> | 0.26 (0.15-0.45) | <.001 | 0.55 (0.25-1.23) | NS |
| Education | Less or equal HSC | Ref |  | Ref |  |
|  | Graduate | 9.07 (6.34-12.97) | <.001 | 6.55 (3.77-11.38) | <.001 |
|  | Master or higher | 5.38 (3.79-7.64) | <.001 | 1.95 (1.16-3.29) | <.01 |
| Income | Less than 30,000 | Ref |  | Ref |  |
|  | 30,000-45,000 | 0.35 (0.22-0.57) | <.001 | 0.38 (0.2-0.74) | <.01 |
|  | 46,000-60,000 | 1.01 (0.67-1.52) | NS | 1.22 (0.64-2.33) | NS |
|  | 61,000-75,000 | 1.41 (1-1.98) | <.05 | 1.25 (0.72-2.18) | NS |
|  | 76,000 and above | 7.13 (4.89-10.39) | <.001 | 9.67 (4.86-19.27) | <.001 |
| Occupation | Service holder (govt/private) | Ref |  | Ref |  |
|  | Entrepreneur/business | 0.78 (0.54-1.14) | NS | 0.68 (0.36-1.28) | NS |
|  | Student | 0.31 (0.23-0.41) | <.001 | 0.19 (0.11-0.32) | <.001 |
|  | Other<sup>y</sup> | 0.35 (0.23-0.52) | <.001 | 0.17 (0.09-0.32) | <.001 |
| Religion | Islam | Ref |  | Ref |  |
|  | Hinduism | 1.74 (1.21-2.5) | <.05 | 1.65 (0.91-2.99) | NS |
|  | Buddhists/Cristian | 1.66 (0.65-4.23) | NS | 3.14 (0.46-21.69) | NS |
| Region | Urban | Ref |  | Ref |  |
|  | Rural | 0.61 (0.49-0.77) | <.001 | 0.46 (0.3-0.69) | <.001 |

NS, not significant; Other<sup>y</sup> includes Housewife, Retired, Unemployed; Other<sup>f</sup> includes Divorced, Widowed, and Separated

**Figure 4:**
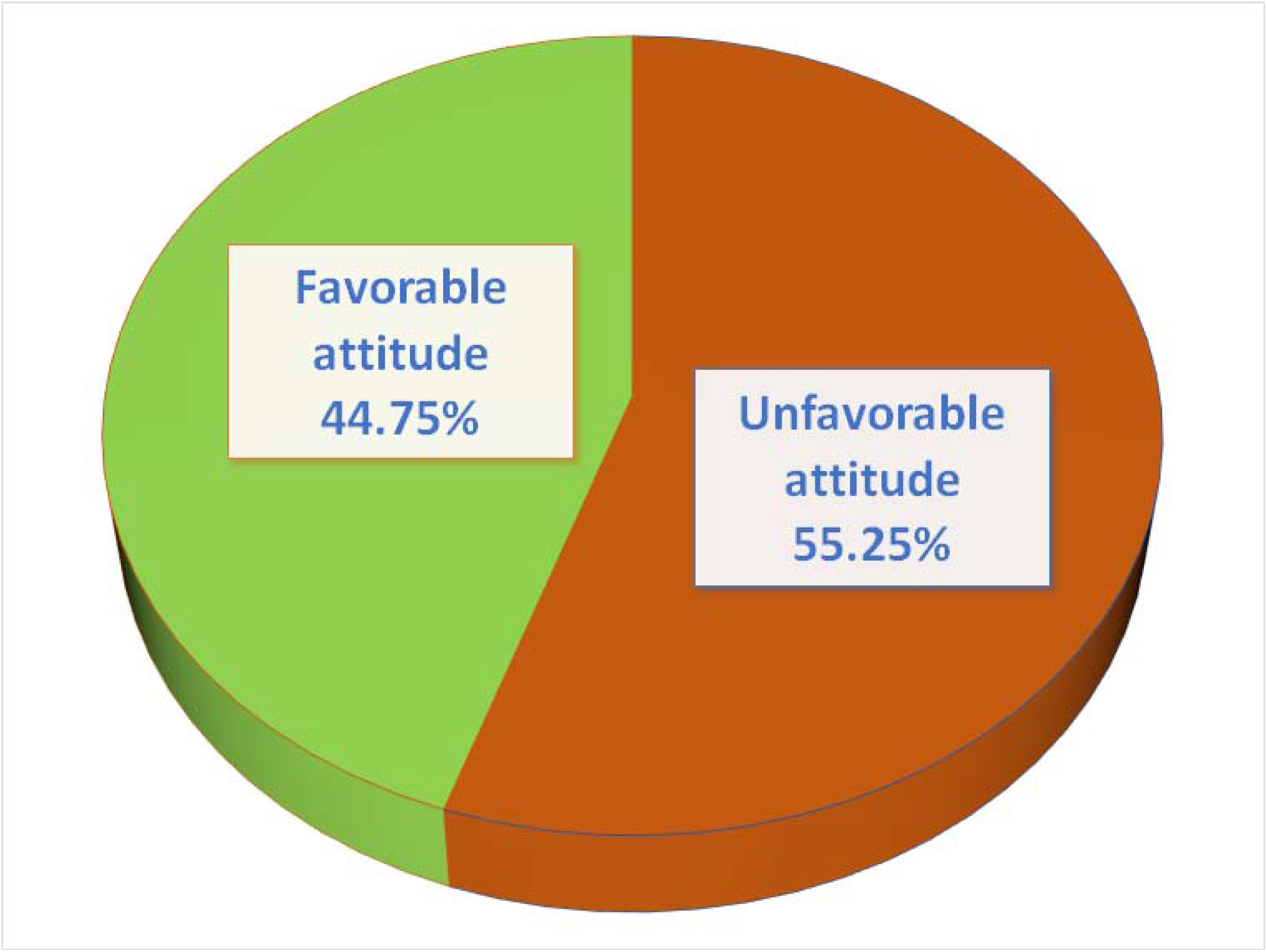
Distribution of attitude towards TB among the general population.

## 4 Discussion

There was a fair blend of male and female participants in the study (male-58.64%). About 80% of the study participants fell in the age category of 15-44 years, which indicates a young study population. However, this age group is of immense interest because, according to Bangladesh’s national tuberculosis program, three-quarters of TB cases in Bangladesh were in the age bracket of 15–45 years [13]. Only 28.14% of study participants had HSC or less (<=12^th^ grade) education level, and a substantial portion of the participants were current students (44.75%). The above socio-demographic characteristics are understandable due to the nature of the survey. We circulated the survey link through social media and other electronic platforms. Young, educated, and student populations understandably have greater access to social media and electronic platforms. Participants also showed a good level of urban-rural balance (rural-56%).

Almost all survey participants in this study had heard of tuberculosis, which is consistent with research undertaken in Nigeria, India, Pakistan, and Lesotho [1,26–28]. The primary sources of information about TB were TV/Radio/Newspapers, Leaflets/Posters/Signboards/Billboards, health professionals, the internet, teachers/religious leaders, family/friends/relatives. The sources of information found in this study are parallel with a previous study conducted in Bangladesh [13]. We found that just 47.8% of respondents possessed sufficient overall knowledge of tuberculosis. A sizable number (71%) of participants knew that tuberculosis germs or bacteria are primarily responsible for TB infection. However, this knowledge level is lower than similar studies conducted in Malawi, Ethiopia, and India (90%, 81.7%, and 81%, respectively). In addition, less than half of the respondents (42%) were aware of the correct mode of transmission of tuberculosis (TB can be transmitted from person to person via coughing or sneezing). This indicates a substantially low level of knowledge about TB in Bangladesh compared to the findings of other studies [1].

The participants’ knowledge regarding signs of TB was insufficient. Less than half of the respondents knew the common symptoms of TB infection (cough for more than three weeks, persistent fever, nighttime sweating, and weight loss). This finding is consistent with similar studies conducted in Bangladesh and Lesotho [1,29]. On the other hand, respondents were well informed about the availability of tuberculosis treatment, with more than three-fourths of respondents being aware of the curability of the disease and the availability of free treatment in Bangladesh. This is consistent with other global studies in Brazil, India, and Tanzania [30–32]. Females had a significantly lower likelihood of possessing adequate knowledge on TB, and middle-aged (30-40 years) people had a significantly higher likelihood of having adequate knowledge than younger and older people. These two results are coherent with a nationwide study conducted in Bangladesh [33]. Higher-income respondents were more likely to have adequate knowledge about tuberculosis, while rural residents had a considerably lower chance of having an adequate understanding of tuberculosis.

Less than a third (31.19%) of 1,180 study participants demonstrated overall good practices (Figure 3). This is an abysmal level of good practices even compared to the estimate of slum dwellers in Nigeria (48.8%) [34] and some other studies in Gambia, Pakistan [35,36]. Gender, age, marital status, education, and religion were significantly associated with good practices towards TB with females, married, younger and older, less educated, and Muslim people showed poor practices towards TB. Although poor practices were found more prevalent in this study than many others, the risk factors for poor practices discovered in this study are consistent with other studies [35–37].

Fewer than half of the participants expressed a favorable attitude about tuberculosis (44.75%) (Figure 4). Lack of proper knowledge about TB might be why this study found such a low level of positive attitudes towards TB. The estimate of favorable attitude is inconsistent with other identical studies showing a higher positive attitude towards TB than this study [30,34–37]. A considerable proportion of respondents wanted to keep it secret if any family member gets TB, were unwilling to work with one previously treated for TB and had a stigmatizing thought about TB patients. Like knowledge and practices, favorable attitudes had similar risk factors. Gender, age, education level, marital status, region (urban/rural) were significantly associated with favorable attitudes towards TB. Unmarried and undergraduate/graduate level respondents displayed 3 and 2-6 times higher likelihood of having favorable attitudes towards TB, respectively. Females and rural people showed a 95% and 54% lower likelihood of possessing positive attitudes, respectively. Covariates identified for attitudes towards TB are accordant with other studies conducted in developing countries like Bangladesh [1,13,38,39]. However, Luba et al. and some other studies found higher positive attitudes among females and married people, which is inconsistent with this study [1,13].

### 4.1 Strengths and Limitations of the study

One of the strengths of our study is that to our knowledge, for the first time in Bangladesh, we conducted a study to assess the knowledge, attitudes, and practices towards TB and their associated factors among general people. Also, our sample size was significantly greater than some other relevant studies. However, this study has some limitations: One of the critical limitations of this research is that the data were collected by sharing an online survey link through social media. Therefore, only individuals with access to social media and the internet took part in the study. As a result, information bias may occur since we collected self-reported data.

## 5 Conclusion

This study revealed poor knowledge, attitudes, and practices towards TB in Bangladesh. Less than half of the study participants showed sufficient knowledge, good practices, and favorable attitudes towards TB. These findings are poorer than most other study findings conducted in developing country settings. Females, older, illiterate/less educated, married, and rural people were more vulnerable to having poor knowledge, attitudes, and practices towards TB. TB is still highly prevalent in Bangladesh. Bangladesh government and policymakers should design programs and interventions to improve knowledge, attitudes, and practices about TB among the general people in a bid to achieve the End TB goals, including a 95% reduction in TB mortality and a 90% reduction in TB incidence by 2035 compared to 2015 levels. More studies are also required to explore other aspects to fight against the TB crisis.

## Data Availability

The data that support the findings of this study are openly available at an Open Science Framework https://doi.org/10.17605/OSF.IO/A9MR2

## Acknowledgments

We are grateful to all who spent their valuable time participating in the survey voluntarily and sharing the link with others. We are also grateful to those who, despite not being eligible, shared the link and inspired others to participate.

## Author Contributions

**SM** initiated and designed the project, designed data collection tools, monitored data collection, wrote the statistical analysis plan, cleaned and analyzed the data, and drafted and revised the paper. MM conceptualized the study, oversaw data collection, conducted data analysis, and wrote and revised the article. SH-I handled data collection, data analysis, and manuscript revision. AM was responsible for supervising data collection and editing the draft manuscript. AI was responsible for coordinating data collection and revising the draft manuscript.

## Data Availability

The data that support the findings of this study are openly available at an Open Science Framework [40].

## Appendix A

**Table A1:** Association of different socio-demographic factors with knowledge of TB, practice and attitude towards TB among the general population of Bangladesh.

| Variables | Adequate knowledge<br>n<br>(%) | Poor knowledge<br>n<br>(%) | χ<sup>2</sup><br>(P-Value) | Good practice<br>n<br>(%) | Bad practice<br>n<br>(%) | χ<sup>2</sup><br>(P-Value) | Favorable Attitude<br>n<br>(%) | Unfavorable Attitude<br>N<br>(%) | χ<sup>2</sup><br>(P-Value) |
| --- | --- | --- | --- | --- | --- | --- | --- | --- | --- |
| Total | 564<br>(47.8) | 616<br>( 52.20) |  | 368<br>(31.19) | 812<br>( 68.81) |  | 528<br>(44.75) | 652<br>(55.25) |  |
| Gender |  |  |  |  |  |  |  |  |  |
| Male | 472<br>(68.21) | 220<br>( 31.79) | 279.40<br>($<.001$) | 260<br>(37.57) | 432<br>(62.43) | 31.80<br>($<.001$) | 464<br>(67.05) | 228<br>(32.95) | 336.74<br>($<.001$) |
| Female | 92<br>(18.85) | 396<br>( 81.15) |  | 108<br>(22.13) | 380<br>(77.87) |  | 64<br>(13.11) | 424<br>(86.89) |  |
| Age |  |  |  |  |  |  |  |  |  |
| 15-29 | 184<br>(45.54) | 220<br>(54.46) | 141.34<br>($<.001$) | 112<br>(27.72) | 292<br>(72.28) | 25.50<br>($<.001$) | 152<br>(37.62) | 252<br>(62.38) | 165.70<br>($<.001$) |
| 30-44 | 332<br>(63.85) | 188<br>(36.15) |  | 200<br>(38.46) | 320<br>(61.54) |  | 332<br>(63.85) | 188<br>(36.15) |  |
| 45-59 | 44<br>(18.33) | 196<br>(81.67) |  | 52<br>(21.67) | 188<br>(78.33) |  | 44<br>(18.33) | 196<br>(81.67) |  |
| 75+ | 4<br>(25.00) | 12<br>(75.00) |  | 4<br>(25.00) | 12<br>(75.00) |  | 0<br>(0.00) | 16<br>(100.00) |  |
| Marital Status |  |  |  |  |  |  |  |  |  |
| Married | 308<br>(41.85) | 428<br>( 58.15) | 189.90<br>($<.001$) | 208<br>(28.26) | 528<br>(71.74) | 25.03<br>($<.001$) | 280<br>(38.04) | 456<br>(61.96) | 147.91<br>($<.001$) |
| Unmarried | 248<br>(75.61) | 80<br>(24.39) |  | 136<br>(41.46) | 192<br>( 58.54) |  | 232<br>(70.73) | 96<br>(29.27) |  |
| Other<sup>£</sup> | 8<br>(6.90) | 108<br>( 93.10) |  | 24<br>(20.69) | 92<br>(79.31) |  | 16<br>(13.79) | 100<br>(86.21) |  |
| Education |  |  |  |  |  |  |  |  |  |
| Less or equal HSC (≤grade 12) | 52<br>(15.66) | 280<br>(84.34) | 192.95<br>($<.001$) | 72<br>(21.69) | 260<br>(78.31) | 26.89<br>($<.001$) | 52<br>(15.66) | 280<br>( 84.34) | 171.96<br>($<.001$) |
| Undergraduate | 256<br>(62.75) | 152<br>(37.25) |  | 124<br>(30.39) | 284<br>(69.61) |  | 256<br>(62.75) | 152<br>( 37.25) |  |
| Master's or higher (Graduate) | 256<br>(58.18) | 184<br>( 41.82) |  | 172<br>(30.09) | 268<br>(60.91) |  | 220<br>(50.00) | 220<br>( 50.00) |  |
| Income |  |  |  |  |  |  |  |  |  |
| Less than 30,000 | 108<br>(39.13) | 168<br>( 60.87) | 227.16<br>($<.001$) | 92<br>(33.33) | 184<br>( 66.67) | 34.89<br>($<.001$) | 96<br>(34.78) | 180<br>(65.22) | 214.84<br>($<.001$) |
| 30,000-45,000 | 36<br>(20.45) | 140<br>( 79.55) |  | 24<br>(13.64) | 152<br>( 86.36) |  | 28<br>(15.91) | 148<br>(84.09) |  |
| 46,000-60,000 | 48<br>(30.00) | 112<br>(70.00) |  | 44<br>(27.50) | 116<br>( 72.50) |  | 56<br>(35.00) | 104<br>(65.00) |  |
| 61,000-75,000 | 132<br>(47.14) | 148<br>( 52.86) |  | 100<br>(35.71) | 180<br>( 64.29) |  | 120<br>(42.86) | 160<br>(57.14) |  |
| 76,000 and above | 240<br>(83.33) | 48<br>(16.67) |  | 108<br>(37.50) | 180<br>(62.50) |  | 228<br>(79.17) | 60<br>(20.83) |  |
| Occupation |  |  |  |  |  |  |  |  |  |
| Service holder (govt/private) | 220<br>(67.90) | 104<br>( 32.10) | 123.12<br>($<.001$) | 104<br>(32.10) | 220<br>( 67.90) | 10.68<br>($<0.05$) | 200<br>(61.73) | 124<br>( 38.27) | 79.07<br>($<.001$) |
| Entrepreneur/business | 108<br>(62.79) | 64<br>( 37.21) |  | 68<br>(39.53) | 104<br>(60.47) |  | 96<br>(55.81) | 76<br>(44.19) |  |
| Student | 168<br>(31.82) | 360<br>(68.18) |  | 160<br>(30.30) | 368<br>(69.70) |  | 176<br>(33.33) | 352<br>(66.67) |  |
| Other<sup>¥</sup> | 68<br>(43.59) | 88<br>(56.41) |  | 36<br>(23.08) | 120<br>(976.92) |  | 56<br>(35.90) | 100<br>(64.10) |  |
| Religion |  |  |  |  |  |  |  |  |  |
| Islam | 468<br>(45.53) | 560<br>(54.47) | 16.97<br>($<.001$) | 290<br>(28.21) | 738<br>(71.79) | 41.00<br>($<.001$) | 442<br>(43.00) | 586<br>(57.00) | 9.89<br>($<0.01$) |
| Hinduism | 86<br>(64.18) | 48<br>(35.82) |  | 74<br>(55.22) | 60<br>(44.78) |  | 76<br>(56.72) | 58<br>(43.28) |  |
| Buddhists/Cristian | 10<br>(55.56) | 8<br>( 44.44) |  | 4<br>(22.22) | 14<br>(77.78) |  | 10<br>(55.56) | 8<br>(44.44) |  |
| Region |  |  |  |  |  |  |  |  |  |
| Urban | 284<br>(55.47) | 228<br>( 44.53) | 21.34<br>($<.001$) | 164<br>(32.03) | 348<br>( 67.97) | 0.30<br>(NS) | 264<br>(51.56) | 248<br>(48.44) | 17.00<br>($<.001$) |
| Rural | 280<br>(41.92) | 388<br>( 58.08) |  | 204<br>(30.54) | 464<br>( 69.46) |  | 264<br>(39.52) | 404<br>(60.48) |  |

NS, not significant; Other<sup>z</sup> includes Housewife, Retired, Unemployed; Other<sup>f</sup> includes Divorced, Widowed, and Separated

